# The Role of ACT Score in Mepolizumab Discontinuation

**DOI:** 10.1101/2023.09.28.23296286

**Authors:** Neha Solanki, Brittany Beck, Monica Labadia, Kevin Smith, Laura Peterson, Stephanie King, Sarah Micklewright, Emily Pennington, Sobia Farooq, Peng Zhang, Mark Aronica, Joe Zein, Sumita Khatri, Suzy Comhair, Serpil Erzurum

## Abstract

**Background:** Mepolizumab is a therapy for severe asthma. However, studies on mepolizumab with external validity and diverse population from the US are lacking. There is little knowledge of the characteristics of people that discontinue mepolizumab in clinical care.

**Objective:** To investigate the real-world efficacy and time to clinical discontinuation of mepolizumab, we evaluated individuals with asthma started on mepolizumab at the Cleveland Clinic. We hypothesized that there are characteristics that distinguish which patients would remain on mepolizumab and which patients would discontinue the medication.

**Methods:** Between 2016 and 2022, patients who started on mepolizumab consented to be assessed over 18 months. At baseline, a questionnaire including demographic and medical history was collected. Laboratory findings such as ACT score, F_E_NO (Fractional Excretion of Nitric Oxide), and spirometry were recorded. At the conclusion of the observation period, the participants were divided into two categories: Group A and Group B.

**Results:** Group B [N=28] discontinued mepolizumab (*P* < 0.05) at an average of 5.8 months (SD 4.2 months). Group A [N=129] stayed on the therapy for at least 1 year. A participant with an ACT score less than 13 has an odds ratio of 6.64 (95% CI, 2.1 – 26.0) of discontinuing mepolizumab therapy. For a male, the odds of discontinuing mepolizumab therapy is 3.39 (95% CI, 1.1 – 11.2).

**Conclusion:** In this real-world study, we find that high eosinophil count may not be adequate in screening which individuals will benefit from mepolizumab. Up to 17% of patients fail therapy within 6 months, with male sex and low ACT score increasing risk of mepolizumab discontinuation at Cleveland Clinic.

## Introduction

People with severe and uncontrolled asthma experience the highest impact on quality of life and accompanying morbidity from asthma. Severe asthma, which is uncontrolled asthma despite adherence to high dose inhaled corticosteroids (ICS) and long-acting beta agonist treatment (LABA), is present in 5%-10% of people with asthma (1, 2). Caregivers have new and exciting management options for steroid-sparing medications including treatment with monoclonal antibodies also known as biologic agents that target immunoglobulin E (IgE), interleukin 5, interleukin-5 receptor, or interleukin-4 receptor (3). The introduction of biologic therapy to the treatment of severe asthma represents an opportunity to carry out phenotype-specific intervention, reinforcing the need for precision in diagnosis (1). Though the Global Initiative for Asthma (GINA) does include a decision tree for how to choose a biologic agent for an individual with severe asthma, many individuals remain uncontrolled and can remain on a biologic agent for longer than necessary (2). People with severe and uncontrolled asthma experience the highest impact on quality of life and accompanying morbidity from asthma. Additionally, according to GINA, the cost of treating individuals with severe asthma is higher when compared to the cost of treating the general population for asthma (2, 4), which further underscores the possible benefits for precision medicine in asthma control and healthcare costs.

GlaxoSmithKline’s (GSK) mepolizumab, a humanized, interleukin-5 antagonist monoclonal antibody, received approval from the US FDA (Federal Drug Administration) in 2015 for add-on treatment in adults with severe eosinophilic asthma (5). The clinical trials that evaluated mepolizumab in patients with asthma included three double-blind, randomized, placebo-controlled studies, including DREAM (Dose Ranging Efficacy And safety with Mepolizumab in severe asthma; NCT01000506), the Phase III MENSA (MEpolizumab as adjuNctive therapy in patients with Severe Asthma; NCT01691521) trial, and SIRIUS (SteroId ReductIon with mepolizUmab Study; NCT01691508) (5). The DREAM trial showed that a baseline blood eosinophil count and increased exacerbation frequency in the previous year were associated with good response to mepolizumab. The confirmatory trials required patients to have a blood eosinophil level greater than or equal to 150 cells/μL at screening or blood eosinophil levels greater than or equal to 300 cells/μL within one year of enrollment (6). SIRIUS, a 24-week clinical trial, demonstrated that mepolizumab has a significant glucocorticoid-sparing effect compared with placebo (6). In patients with baseline eosinophil counts greater than or equal to 150 cells//μL, the DREAM, MENSA, and MUSCA (Mepolizumab adjUnctive therapy in subjects with Severe eosinophiliC Asthma; NCT02281318) trials demonstrated that mepolizumab treatment reduces exacerbations, increases forced expiratory volume in one second (FEV_1_) and improves asthma control and quality of life (5). While clinical trials are pivotal for FDA-approval of new therapies, real world applications of medicines can often reveal different outcomes than in prospective blinded trials (7).

To assess real-world effects of mepolizumab, Glaxo Smith Kline (GSK) funded REALITI-A (REAL world effectiveness of mepolizumab in patient care – Asthma), a large prospective study with 368 participants. REALITI-A demonstrated a meaningful reduction in daily maintenance oral corticosteroid dose for people with asthma and a decrease in the rate of exacerbations. However, there was not a difference shown for FEV_1_ (forced expiratory value in 1 second) % (5). Observational real-world cohorts from Europe also showed that mepolizumab reduces prednisone dependence and improves asthma control (8-10). Israeli and Australian studies demonstrate an additional FEV_1_% improvement (11, 12). The results from this real-world study suggest that more data are needed regarding the effects of mepolizumab on FEV_1_% and the time to mepolizumab’s clinical failure for a person with very severe asthma.

To further investigate the real-world efficacy of mepolizumab, time to discontinuation of mepolizumab and identification of clinical traits of those who remain on mepolizumab, we evaluated people with asthma treated with mepolizumab at the Asthma Center of Cleveland Clinic over 6 years. We hypothesized that there may be baseline characteristics that distinguish which patients will respond to mepolizumab and the characteristics of the type of response, including better asthma control, lower corticosteroid dose, and better lung functions.

## Methods

### Baseline Characteristics

Cleveland Clinic’s biologic registry includes individuals on omalizumab, mepolizumab, benralizumab, reslizumab, dupilumab, and tezepelumab. An asthma specialist assessed eligible participants in the registry and prescribed mepolizumab based on standard U.S guidelines at the time of study. These guidelines included an absolute blood count eosinophil ≥ 150 cells/ μL or corticosteroid dependence and clinical evidence of uncontrolled asthma as demonstrated by two or more asthma exacerbations requiring prednisone, one or more hospitalizations or emergency medical visits or poor symptom control while being on an inhaled corticosteroid and a long-acting beta-agonist. Between 2016 and 2022, we consented 199 people who were prescribed mepolizumab by their provider. The Institutional Review Board (IRB # 8351) at Cleveland Clinic approved the study protocol and procedure.

### Longitudinal Assessment

The study team assessed each participant at baseline visit prior to first dose of mepolizumab, and then at 3 months, 6 months, 12 months, and 18 months (Figure 1). At the baseline visit, our team collected a questionnaire which included demographic information, asthma history, medical history, review of systems, asthma medications, and healthcare utilization from each participant (Figure 1). We also recorded peak flow, ACT score, F_E_NO, spirometry and CBC, and we conducted similar assessments of the participants in subsequent visits.

### Outcomes

The outcome of this retrospective study was discontinuation of mepolizumab therapy based on lack of clinical response, which was determined by the asthma specialist and informed by the patient’s perspective and the provider’s medical reasoning. After 18 months of serial assessments, participants who started mepolizumab were divided into two categories: Group A and Group B. We defined Group A participants received mepolizumab for at least six months or received mepolizumab for 18 months without a change in therapy. Group B participants stopped the therapy within 6 months or had a clinical need to switch to a different biologic therapy within 18 months. Chart review excluded participants who stopped mepolizumab therapy due to other reasons.

### Statistical analysis

We performed all analyses with R, version 4.1.2 (R Project for Statistical Computing, Vienna, Austria), and we reported results as mean with standard deviation or median with interquartile range depending on whether the value was parametric or non-parametric. For the descriptive analysis, we used student’s t-test or Kruskal-Wallis test to compare participants on mepolizumab to those on other biologics and to compare mepolizumab responders and non-responders. Categorical variables were analyzed using chi-squared tests. Comorbidities that that have 10% or higher missing data were excluded. Several columns that had over 10% of “missing” values were excluded. We used a Kaplan-Meier curve to compare time on the drug for mepolizumab responders and non-responders. Each responders’ time was censored after 18 months. When data were censored, the total time on mepolizumab for a subject cannot be accurately determined past a certain date. Log rank was used to calculate *P* for the Kaplan-Meier curve. A receiver operative curve (ROC) with C-index was used to determine optimal cutoff for discrete variables with *P* < 0.05 between responders and non-responders. For the inferential analysis, binomial logistic regression was utilized to obtain odds ratio with confidence intervals. In this analysis, less than 4% of data were missing and were deemed missing completely at random.

## Results

Of the participants who started on mepolizumab (*n* = 199), the top reason for exclusion is lack of recorded follow-up visits (*n*= 27). Participants (*n* = 15) are excluded for medication side effects, loss to follow up, or change in medication indication (Figure 2a). 157 (70.4%) of the participants who started on mepolizumab at their baseline visits are included in this analysis (Figure 2b). The group that started mepolizumab and the group that started other biologic agents have similar baseline demographic characteristics and medical comorbidities (Table 1) except for a higher prevalence nasal polyps (*n* = 69 versus [vs] *n* = 16, *P* < 0.05) and sinusitis (*n* = 112 vs *n* = 30, *P* < 0.05) in participants placed on mepolizumab. Participants starting mepolizumab are also more prednisone dependent (*n* = 98 vs *n* = 24, *P* < 0.001). There are no differences in asthma control score, eosinophil counts, and daily oral prednisone use between the two groups (Table 1).

For the participants placed on mepolizumab (*n* = 157), there is gradual attrition over visits 3, 6, 12, 18 months (Figure 3). Despite attrition, median ACT scores improve over sequential visits on mepolizumab (*P* < 0.05) (Figure 4-A). FEV_1_ improves rapidly in most individuals between the baseline visit and 3 months, however it is not sustained (*P* < 0.05) (Figure 4-B). There is a striking decline in prednisone use between visit 1 and subsequent visits (*P* < 0.05) (Figure 4-C).

Of the 157 individuals on mepolizumab, 129 are classified as Group A and 28 as Group B (Table 2). Group B participants are on mepolizumab therapy (*P* < 0.05) for an average of 5.8 months (standard deviation [SD] 4.2 months) (Figure 5). Participants in Group A demonstrated a higher ACT score at each subsequent visit (Figure 6A); whereas participants in Group B did not demonstrate a change in ACT score through subsequent visits (Figure 6B).Group B participants have lower median baseline ACT of 9 (interquartile range [IQR], 4) compared to 13 (IQR, 9) for responders (*P* < 0.05) (See Figure 7). There are no other differences between the baseline characteristics of the two groups (Table 2). Sub-analysis of people with nasal polyps demonstrates that there is no difference in the people who discontinue mepolizumab and those who stay on the medication (Table 3).

Based on the observed difference in ACT of the groups, the area under the curve (AUC) of the receiver operative curve analysis was 0.68 for an ACT cutoff score of 13 (Figure 8). Binomial logistic regression controlled for age, BMI (Body Mass Index), F_E_NO, and eosinophils show that for a participant who has an ACT score less than 13, the odds ratio of being in Group B is 6.64 (95% CI, 2.1 – 26.0) (Figure 9). Men have an odds ratio of 3.39 (95% CI, 1.1 – 11.2) for being in Group B (Figure 9)

## Discussion

Utilizing this real-world cohort of people with severe asthma, we find that most people on mepolizumab demonstrate clinical improvement shown by a decrease in ACT score by 18 months, a decrease in prednisone use as soon as 3 months, and an initial increase in FEV_1_% by 3 months. Our findings also corroborate the REALITI-A trial and other real-world studies, but unlike others, this study is sponsored in part by NIH. When compared to the cohorts on other biologic therapy, the mepolizumab cohort comprises this registry’s longest and most complete longitudinal data. This study, which is the largest US real-world mepolizumab to date, shows that despite having eosinophils ≥ 150 cells/μL and history of recurrent asthma exacerbations, male sex or a person with severe asthma who has an ACT score < 13 is more likely to discontinue mepolizumab therapy. Our findings also show that if mepolizumab is discontinued, the discontinuation will occur within 6 months of starting the drug; those that stay on mepolizumab and demonstrate an improvement in ACT score are on mepolizumab for at least an average of 1 year. The results suggest that poorly controlled asthma at baseline and men with severe asthma may have a different biologic endotype less responsive to IL-5 therapy. A sub-analysis of Group B including those individuals initially excluded for side effects demonstrates that sex remains a critical variable for discontinuing mepolizumab at 6 months. Additionally, those with a lower baseline ACT score still are more likely to discontinue mepolizumab at 6 months. These findings in the intention-to-treat sub-analysis bolster the initial findings (Table 4).

Participants of Group B may have a complex neutrophilic or pauci-granulocytic mechanism for their asthma because Type 2 (T2) inflammation-mediated asthma responds well to mepolizumab (13). Another possibility is that their asthma may be not controlled well at baseline. Predictive biomarkers for response to mepolizumab treatments so far include blood eosinophil counts as the strongest predictor for the reduction in exacerbation rates with mepolizumab compared to placebo (14). However, in our study, eosinophil counts are similar among responders and non-responders. The Australian Mepolizumab Registry did find that higher blood eosinophil levels and a better baseline Asthma Control Questionnaire (ACQ-5) predicted a superior clinical response to mepolizumab (11).

Regarding sex as a risk factor, 31% of the participants in our study on mepolizumab are male. While most of the participants in our study are female, males were found to have an odds ratio of 3.39 of failing mepolizumab therapy. The Australian Mepolizumab Registry also found that males were more likely to fail mepolizumab therapy (11). Sex hormones play a role in asthma severity, and androgens such as testosterone may reduce asthma incidence and symptoms (15). Therefore, the finding that males have higher odds of failing mepolizumab therapy is unexpected and may suggest underlying sex-based mechanism for this subset.

For our study, the median FEV_1_% improves by 3 months on the therapy. Most people also have a decrease in prednisone use by the third month. Therefore, we expected FEV_1_% to stay consistent or continue improving after visit 3, but we do not find any remarkable differences past the third month on mepolizumab. Therefore, while there may be an initial improvement in FEV_1_% by the third month, the effect is not long-lasting. Pre-bronchodilator FEV_1_% is lower at baseline for participants (though *P* > 0.05) who are clinical non-responders compared to clinical responders. If the study had higher power, it is possible that this difference would become more pronounced. REALITI-A does not show a difference shown for FEV_1_% (5), but Israeli and Australian studies demonstrate an FEV_1_% improvement (11, 12).Obesity, which causes changes to normal lung physiology, innate immune function, and changes in the gut microbiome, is a major risk factor and disease modifier in asthma for adults (16). The BMI values of people on mepolizumab trend higher than the BMI values of people placed on other biologics (32.1 [9.7] vs 30 [12.1] (*P* = 0.171), and the BMI values of Group B trend higher than the BMI values of Group A (32.5 [7.8] vs 31 [12] (*P* = 0.513). Obesity causes mechanical changes such as lung compression and reduction in lung volume (16). As BMI increases, there is a higher risk for airway hyperreactivity (17). Additionally, the Western dietary pattern, which promotes obesity, might also affect the development of asthma via changes in gut microbiome (16).

While this study has many strengths such as strong external validity because it is a real-world study and has a diverse demographic population, it has three limitations. In a real-world study, there are no controls and no randomization. The first limitation in our study is that participants who started on mepolizumab compared to those that started on other biologic agents have higher a prevalence of chronic rhinosinusitis with nasal polyposis (CRSwNP). After the randomized controlled trial, SYNAPSE, mepolizumab was approved for nasal polyps in the setting of severe chronic rhinosinusitis (18). However, those with greater type 2 inflammation often have concomitant CRSwNP, so it is not surprising that clinicians chose mepolizumab for those with this comorbidity (19). Since the people started on mepolizumab have a higher incidence of CRSwNP in this study, these people likely have greater Th2 inflammation which suggests that the population on mepolizumab, regardless of when they discontinue the medication, are going to have more severe asthma at baseline than the people placed on other biologics, which is supported by the higher use of prednisone (62% vs 36%, P < 0.001). Our sub-analysis of the cohort with CRSwNP demonstrated that there was no difference in the people that stayed on mepolizumab and those that discontinued it early (Table 3).

A second limitation is that the individuals in Group A have either milder asthma or better controlled at baseline as they have a higher baseline ACT score and a better baseline FEV_1_ (72% vs 62% predicted, *P* = 0.07). It is difficult to ascertain just based on this observational review whether the participants of Group B have asthma that is severe and uncontrolled or just uncontrolled at baseline. Finally, since this was a real-world study, there is also attrition between the baseline visit and visit 18, which reduces the power of our study. If an individual left the study to receive their asthma management at another facility, their information was not recorded. This further reduces the power of our study. It is remarkable that given the limitations, we still were able to corroborate the findings of DREAM, MENSA, MUSCA, SIRIUS, and REALITI-A, and we were also able to find a strong relationship between ACT score and mepolizumab response.

In conclusion, upto 17% of patients discontinue therapy within 6 months, with male sex and low ACT score increasing risk of early discontinuation.. One mechanism for low ACT scores in the people that discontinue mepolizumab early could be related to altered gut microbiome in the setting of obesity with an altered immune response favoring low Th2 pathways that would be less responsive to biologics targeting Th2 high pathways, despite having a high baseline eosinophil count. Further studies are needed to determine which, if any, biologics might perform better in those people who suffer with severe asthma and poor control.

## Data Availability

All data produced in the present study are available upon reasonable request to the authors.

## Abbreviations

ACT: asthma control test
AUC: area under curve
BMI: body mass index
CAD: coronary artery disease
CBC: complete blood count
CHF: congestive heart failure
DBP: diastolic blood pressure
FDA: Federal Drug Administration
F_E_NO: fractional excretion of nitric oxide
FEV_1_: forced expiratory volume in one second
GERD: gastroesophageal reflux disease
GINA: Global Initiative for Asthma
GSK: GlaxoSmithKline
ICS: inhaled corticosteroids
IQR: interquartile range
IRB: Institutional Review Board
LABA: Long-acting beta agonist
μL: microliters
ROC: receiver operating curve
SBP: systolic blood pressure
WBC: white blood cell

